# COVID-19 in Japan: insights from the first three months of the epidemic

**DOI:** 10.1101/2022.02.10.22270735

**Authors:** Natsuko Imai, Katy AM Gaythorpe, Sangeeta Bhatia, Tara D Mangal, Gina Cuomo-Dannenburg, H Juliette T Unwin, Elita Jauneikaite, Neil M Ferguson

## Abstract

**Background:** Understanding the characteristics and natural history of novel pathogens is crucial to inform successful control measures. Japan was one of the first affected countries in the COVID-19 pandemic reporting their first case on 14 January 2020. Interventions including airport screening, contact tracing, and cluster investigations were quickly implemented. Here we present insights from the first 3 months of the epidemic in Japan based on detailed case data.

**Methods:** We conducted descriptive analyses based on information systematically extracted from individual case reports from 13 January to 31 March 2020 including patient demographics, date of report and symptom onset, symptom progression, travel history, and contact type. We analysed symptom progression and estimated the time-varying reproduction number, *R*_*t*_, correcting for epidemic growth using an established Bayesian framework. Key delays and the age-specific probability of transmission were estimated using data on exposures and transmission pairs.

**Results:** The corrected fitted mean onset-to-reporting delay after the peak was 4 days (standard deviation: ±2 days). Early transmission was driven primarily by returning travellers with *R*_*t*_ peaking at 2.4 (95%CrI:1.6, 3.3) nationally. In the final week of the trusted period, *R*_*t*_ accounting for importations diverged from overall *R*_*t*_ at 1.1 (95% CrI: 1.0, 1.2) compared to 1.5 (95% CrI: 1.3, 1.6) respectively. Household (39.0%) and workplace (11.6%) exposures were the most frequently reported potential source of infection. The estimated probability of transmission was assortative by age. Across all age groups, cases most frequently onset with cough, fever, and fatigue. There were no reported cases of patients <20 years old developing pneumonia or severe respiratory symptoms.

**Conclusions:** Information collected in the early phases of an outbreak are important in characterising any novel pathogen. Timely recognition of key symptoms and high-risk settings for transmission can help to inform response strategies. The data analysed here were the result of robust and timely investigations and demonstrate the improvements to epidemic control as a result of such surveillance.

## Introduction

Japan’s response to the COVID-19 pandemic in the first three months was successful in preventing an initial severe wave of SARS-CoV-2 transmission despite the absence of stringent lockdown policies adopted in other high-income countries [1]. Emphasis was placed on border control, intensive backward contact-tracing, and identification of clusters [2]. Detailed cluster investigations informed Japan’s “3-Cs” strategy of encouraging individuals to avoid “crowded places”, “close-contact settings”, and “confined and enclosed spaces” that was subsequently adopted by the World Health Organization [3, 4]. When the first “state of emergency” (SOE) order was lifted on 25 May, 16,445 cases and 846 COVID-19 deaths had been reported in Japan [5]. However, community transmission increased after the SOE was lifted and further increased due to the omicron variant. As of 8 February 2022, over 3.3M cases and over 19,000 deaths have been reported nationally [6].

Understanding the characteristics and natural history of novel pathogens is crucial to inform successful control measures. Here we analyse data from the first 3 months of the COVID-19 epidemic in Japan and characterise the transmissibility, high-risk settings and age-dependent probabilities of transmission, and clinical progression.

## Methods

### Data collection

We systematically reviewed case reports published by the Ministry of Health of Japan for the period 14 January (date of the first reported case) to 31 March 2020 [7]. Information on age (decade), sex, and geographic location of the case, dates of report, symptom onset, symptom progression, hospitalisation, death or recovery, and suspected exposure, healthcare seeking behaviour, and travel history were manually extracted. For cases with known epidemiological links, we also collected information on the number and type of contacts.

Information on interventions implemented in Japan were collated from the Oxford Covid-19 Government Response Tracker (OxCGRT) [8] and from a manual search of Ministry of Health announcements and media reports [7]. The two datasets were then cross-checked and collated (see supporting information). Mobility data were extracted from the V-RESAS website which reported the average percentage mobility change compared to 2019 baselines [9].

### Delay distributions

We fitted a discrete gamma distribution to the observed delay from symptom onset to case report. We estimated the peak of the observed symptom onset dates by multinomial bootstrapping sampling with replacement using the R package *incidence* [10], and estimated the growth or decay rate before and after this peak. As shorter delays are more likely to be observed during a growing epidemic, this can bias naïve estimates of delay distributions [11, 12]. We therefore corrected the fitted distribution accounting for the growth and decline of the epidemic before and after this peak.

Due to the delay in reporting of cases, the observed incidence is necessarily incomplete. We used the delay from symptom onset to report to identify a trusted period defined as the date by which 95% of the cases have been reported. We use the incidence in the trusted period to estimate the time-varying reproduction number, *R*_*t*_.

### Time-varying reproduction number, R_t_

We quantified the transmissibility as measured by the reproduction number (*R*_*t*_, the average number of secondary cases infected by one individual) using an established approach [13– 15] implemented using the R package *EpiEstim* [16] from case incidence by date of onset over a 7-day sliding window. We assumed a gamma distributed serial interval (time between symptom onset in a case and their infector in a chain of transmission) with mean 7.8 days and standard deviation 5.2 days [17]. We assumed that the daily incidence can be approximated by a Poisson process using the renewal equation:

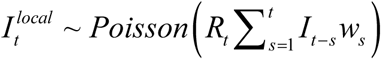

Where 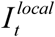 is the incidence of locally acquired cases on day *t, R*_*t*_ the reproduction number on day *t,I*_*t* − *s*_ is the total incidence cases on day *t – s*, and *w* is the probability mass function of the serial interval. We truncated the incidence curve using the trusted period defined above to account for the symptom onset to reporting delay. *R*_*t*_ was only estimated over the trusted period. As a sensitivity analysis, we also used the naïve fitted onset-to-reporting delay to truncate the incidence and estimate *R*_*t*_.

We accounted for imported cases which can bias the estimation of *R*_*t*_ [18] and compared this to estimates assuming all reported cases were locally acquired.

### Contact types over time

Exposure types were aggregated from 32 unique initial categories into 8 broad categories; i) care facility; ii) cruise ship; iii) health care; iv) household or family; v) live music venues; vi) sport facilities; vii) tourism; and viii) workplace or education. We estimated the change in the cumulative proportion of cases with known and unknown exposure types attributed to these 8 categories over time.

### Age-specific transmission matrix

We estimated the age-specific transmission matrix using transmission pairs where a case was linked to a known infector(s). Where multiple possible infectors were reported, one entry was included for each linked pair. The infector case was defined as the source of infection and the secondary case was the infectee. Linked transmission pairs were defined as pairs of COVID-19 cases with known contact within a specified timeframe (≤ 10 days). Any pairs with missing data on age or date of symptom onset were excluded. Serial intervals between the linked pairs were calculated and pairs with negative serial intervals greater than ten days were excluded. A bootstrap approach, using random sampling with replacement, was used to estimate the probability of linked transmission pairs of each age-group. From the full line-list of linked transmission pairs, *N* samples were drawn (where *N* is the number of all linked pairs) and the probability of transmission occurring between age-groups *i* and *j* was calculated as follows:

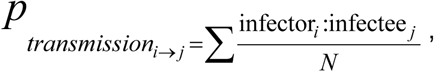

where the numerator describes the frequency of all transmission events from an infector in age-group *i* to an infectee in age-group *j* and is divided by N. This was repeated 1000 times, and the median, 2.5^th^ and 97.5^th^ quantiles are presented.

### Symptom progression

We examined the progression of symptoms after reported onset. Symptoms were classified into common groups, for example, “difficulty breathing” or “breathlessness” were both classified as “respiratory symptoms”. The first date that each symptom type was reported was extracted from the case reports to produce a distribution of time to first report of each symptom group for each decadal age group. To assess for significant differences between the progression of symptoms in each age group, we performed a two-sample Kolmogorov-Smirnov test [19, 20]. This tests the hypothesis that both samples, in this case the samples of time from reported onset of any symptom to first occurrence of a specific symptom for each age group, are drawn from the same continuous distribution. We reject this hypothesis if the p-value of the statistic is below 0.05. For some individuals, a particular symptom group may not be reported at any stage. To emphasise that this symptom progression was different, we set the symptom onset time for these individuals by sampling from a normal distribution with mean -100 and standard deviation 1. Note that in age groups with few individuals, there may be insufficient samples to produce a significant result and thus fail to reject the hypothesis that the samples are drawn from the same distribution.

All analyses were conducted in R version 3.6.3 [21].

## Results

### Demographics, travel history, and exposure types

A total of 2,116 cases of PCR-confirmed SARS-CoV-2 were reported in Japan between 14 January and 31 March 2020. 1,487 cases had a reported date of symptom onset ranging from 3 January to 31 March 2020. 178 individuals were asymptomatic at the time of testing positive, and 15 cases were repatriated from Wuhan City, China. 247 cases had a recent history of international travel, and 1,617 cases were locally acquired. Travel history was unknown for 252 cases. Of the imported cases, the majority (n=151) were from Europe, 28 were from USA, and 26 were from China. Of the cases with reported sex, 58.2% (n=1101/1,893) were male. Age was reported to the nearest decile with the highest number of cases reported in the 50-to 60-year-old age group (n=338, Additional File 1, Figure 1). By 31 March 2020, 45 of 47 prefectures had confirmed cases of COVID-19 with Tokyo (n=485), Osaka (n=227), Hokkaido (n=172), Aichi (n=155), and Hyogo (n=140) the top five affected prefectures. 38% of cases (n=798) had a known contact or contact type with a SARS-CoV-2 PCR-confirmed case. Household and/or family contacts were the most frequently reported close contact type (39.1%), followed by care homes (21.6%) and workplaces (11.6%) (Additional File 1, Figure 2).

**Figure 1:**
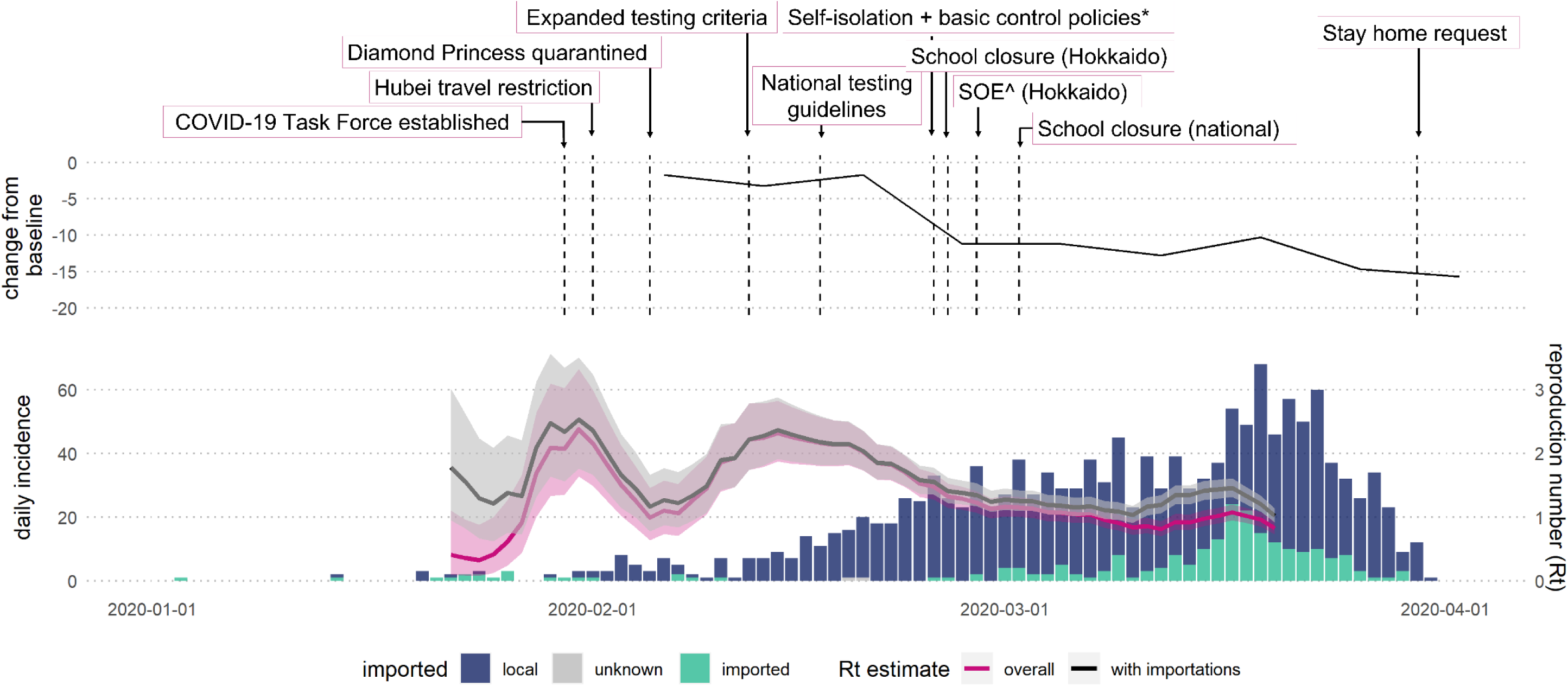
Daily incidence of confirmed COVID-19 cases by date of symptom onset and the estimated time-varying reproduction number (Rt): overall (pink) and accounting for imported cases (black). The solid line shows the median and the shaded area the 95% credible interval. Top panel show the timing of key intervention and the change in mobility compared to 2019. *Basic control policies include: avoiding high risk settings; observing cough etiquette, and wearing a face covering [2].

**Figure 2:**
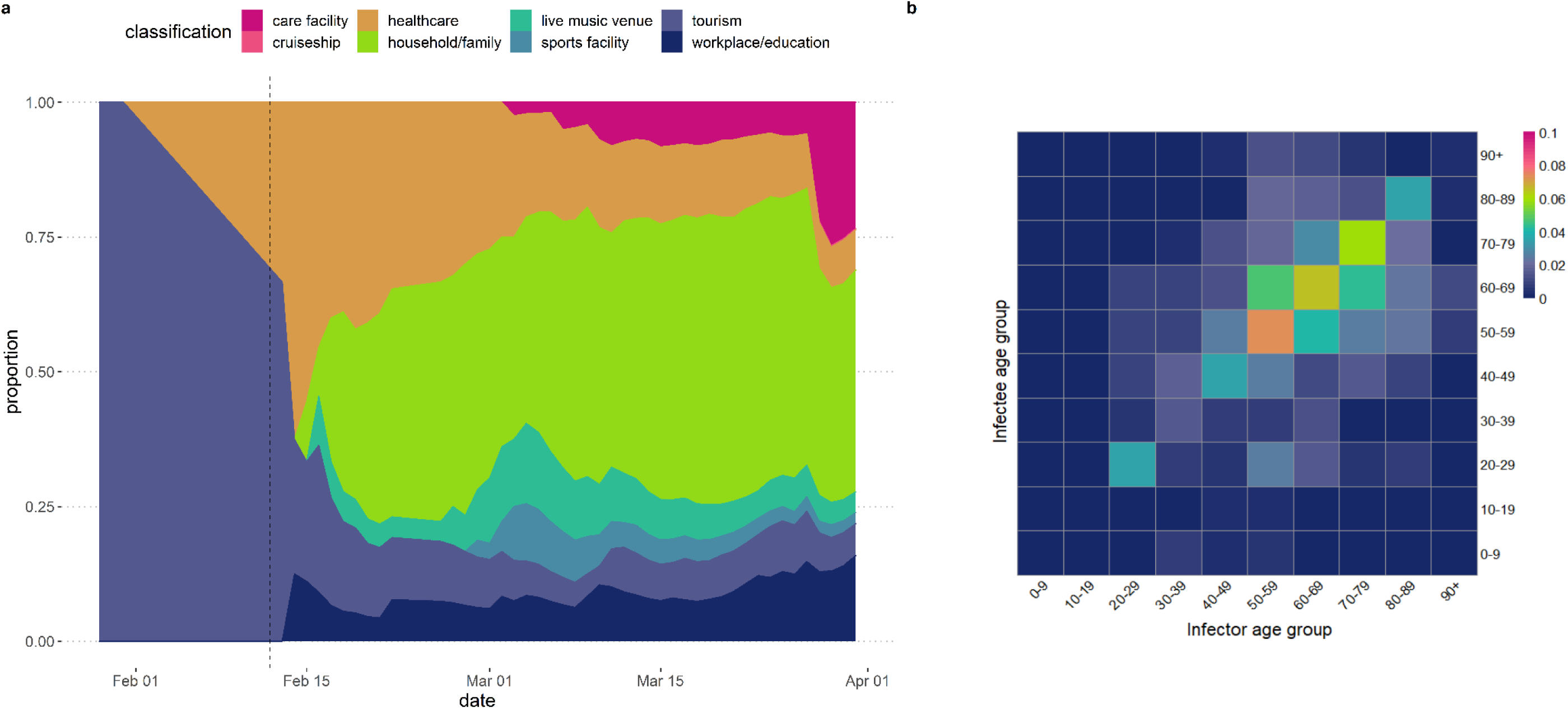
(A) Change in known contact exposure type over time by date of report. The vertical dashed line denotes when the testing criteria was expanded. (B) Age-specific transmission probability matrix. The colours represent the probability of infector-infectee transmission pairs in each 10-year age-group amongst cases reported up to 31 March 2020 in Japan.

### Interventions and Transmission

Japan quickly designated SARS-CoV-2 as a quarantinable infectious disease and established the COVID-19 Task Force two-weeks after the first reported case. Travel restrictions to and from Hubei and Zheijang provinces were implemented on 1 February, and testing was expanded on 12 February to include anyone with respiratory symptoms or a fever regardless of travel history. From 25 February a series of interventions [2] were introduced including self-isolation if symptomatic, requested suspension of large-scale gatherings, information campaigns for hygiene measures, and the establishment of the Cluster Response Team (Additional File 1, Supplementary Table 1). Although none of these measures were legislated, national school closures on 2 March led to a gradual decline in population mobility to 15% lower than 2019 in the equivalent week by the end of March 2020 (Figure 1).

The estimated peak of the observed cases by symptom onset between 3 January and 31 March 2020 was 19 March 2020 (95% CI: 17 March, 23 March). The estimated growth or decline rate before and after the peak was 0.06 (95% CI: 0.06, 0.07) and -0.24 (95% CI: - 0.35, -0.12) respectively. These rates correspond to a doubling time of 11.06 days (95% CI 9.87-12.57 days) and halving time of 2.92 days (95% CI 1.98-5.57 days) respectively. Adjusting for this decline, the mean fitted onset to reporting delay after the peak was 4 days (standard deviation: ±2 days) corrected from 5 days (sd: ±2 days). Based on this adjusted distribution (Additional File 1, Supplementary Figure 3) we estimated that 95% of cases that had symptom onset by 23 March would have been reported by 31 March 2020 and estimate *R*_*t*_ up to this date. Figure 1 shows how estimates of *R*_*t*_ varied over time within this trusted period. *R*_*t*_ peaked twice, on 31 January at 2.4 (95% CrI: 1.6, 3.3) then again on 14 February at 2.3 (95% CrI: 1.9, 2.8). Up to this point, the overall *R*_*t*_ did not differ significantly from *R*_*t*_ accounting for imported cases. However, in the final week *R*_*t*_ accounting for importations diverged from overall *R*_*t*_ at 1.1 (95% CrI: 1.0, 1.2) compared to 1.5 (95% CrI: 1.3, 1.6) respectively.

### Exposures and age-specific transmission matrix

Of 2,116 reported cases in the first wave, 563 had been linked to at least one primary COVID-19 case. Figure 2A shows early cases with known exposures were associated with travel (tourists visiting from Wuhan City, China) and any health care settings. The types of exposures then increased after the testing criteria was expanded on 12 February. The proportion of cases with unknown exposure types remained relatively constant over time (Additional File 1, Supplementary Figure 4) with household or family exposures being the most frequently reported contact type of the known exposures (Figure 2A).

In total, 947 linked transmission pairs were identified (including multiple contacts or clusters for some cases), with the majority between ages 50- and 80-years. Of these, 86 were missing contact age data, 70 were missing contact onset date, 154 were duplicates pairs, and 17 had large negative serial intervals (≥ 10 days), resulting in 620 retained linked pairs. The empirical median serial interval was 1 day (mode: 0 days, range: -10 to 21 days). Figure 2B shows age-assortative patterns with the highest probability of transmission amongst individuals aged 50-59 years and 60-69 years reflecting the age distribution of reported cases.

### Symptom progression

The most common symptom at symptom onset across all age groups was cough, fatigue, or fever (Figure 3). Older age groups, 50 years and above, reported pneumonia or other severe respiratory symptoms earlier in their symptom progression compared to younger age groups. None of the 33 cases aged 0-19 years with known symptom progression reported pneumonia or breathing difficulties suggesting a milder clinical course. Additional File 1, Supplementary table 3 reports the significance of the Kolmogorov-Smirnov test for each significant pair of age groups in terms of symptom occurrence. The onset of pneumonia varies most significantly between youngest and oldest age groups. We also estimate significant differences in the timing of the onset of cough between 20-29 years and 60-69 or 80+ years and between 50-59 years and 80+ years and the onset of respiratory symptoms between 50-59 and 80+ years. There were no significant differences in the timing of the onset of fever between age groups. These results suggest that symptom appearance and occurrence vary between age groups.

**Figure 3:**
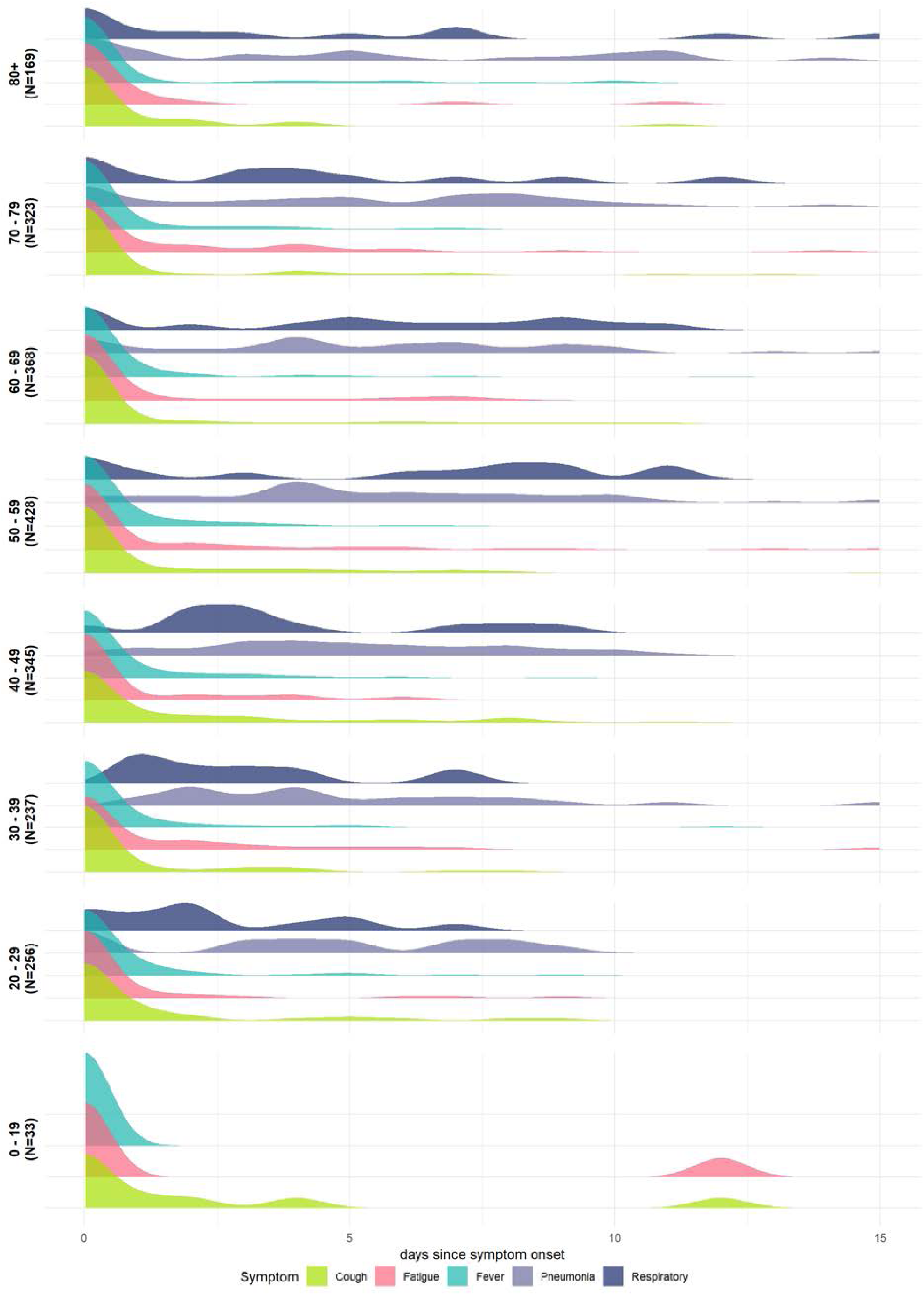
Distribution of symptom progression in days since symptom onset by 10-year age group. N presents the number of people in each age group.

## Discussion

This analysis of the first three months of the COVID-19 epidemic in Japan highlights how detailed outbreak investigations can yield valuable information about transmissibility, at-risk settings or exposure types, and clinical progression relevant for refining case definitions of a novel pathogen. Japan’s early response to COVID-19 was unique in pursuing an intensive cluster-based approach of backward contact-tracing without strict lockdown measures [22, 23]. Although measures could not be enforced, the government requested people to cancel non-essential outings [2] which combined with school closures was estimated to have significantly reduced transmissibility [24]. A 19-fold greater odds of transmission in a closed environment compared to well ventilated settings was estimated from rapid analysis of early clusters [3, 22]. This informed Japan’s “3-Cs” strategy of encouraging individuals to avoid “crowded places”, “close-contact settings”, and “confined and enclosed spaces” that was subsequently adopted by the World Health Organization [3, 4].

We found that initial transmission was driven by travellers from mainland China followed by more localised transmission. However, the surge in case numbers in late March, when overall *R*_*t*_ and *R*_*t*_ accounting for importations diverged (Figure 1), was due to returning travellers from Europe (49.7% of imported cases with known origin were from Europe) where national lockdowns were implemented in Italy, Spain, and the UK on the 11, 14, 24 March 2020 respectively [8, 25]. Other studies have also found a strong positive correlation between the case numbers at prefecture level and the number of international travellers, with SARS-CoV-2 cases detected before March belonging to the Chinese lineages and to the European and American lineages after March 2020 [26]. Mobility gradually decreased from late February, plateauing at minus 10% relative to 2019 levels with a corresponding decline in transmissibility (Figure 1). Our *R*_*t*_ estimate assumes that reporting rates stay constant.

Although PCR-testing increased in March (Additional File 1, Figure 5) [6], this was a period when our estimated *Rt* was declining (Figure 1). Therefore, the decreasing *R*_*t*_ after the second peak in mid-February is likely a true decline. Studies using detailed mobility data suggest that mobility was already decreasing in Tokyo, Kanagawa, Saitama, Chiba, Osaka, Hyogo, and Fukuoka prefectures before the first state of emergency on 7 April with the degree of reduction differing by age group [27], region, and type of establishment [28].

Adams et al. found that social settings were associated with younger and more secondary cases than household settings when controlling for age. Our results are consistent with their observation that households were the most the frequently (54.4%) identified transmission settings [29]. However, household contacts are easier to recall, trace, and test than social or workplace contacts. The change in known contact types over time (Figure 2A) reflects this cluster-based approach with several large clusters identified from live music venues as well as health care settings. Early in the epidemic, hospital-based clusters were frequently reported with over 60 hospitals affected by mid-April [30]. Up to 31 March 2020, we estimate that the proportion of cases with unknown exposure types remained relatively constant over time (Additional File 1, Supplementary Figure 4A). However, in late March, the National COVID-19 Task Force highlighted the increasing proportion of cases across multiple prefectures that were not epidemiologically linked and warned that broader “lockdown type” control measures in addition to the existing cluster-based approach may be required to prevent an explosive growth in case numbers [31].

Detailed investigations of COVID-19 clusters (≥5 cases with a common primary exposure) found that the most probable primary case of cluster outbreaks were 20-39 years old with 41% of probable primary cases being pre-or asymptomatic at time of transmission [22].

Amongst known transmission pairs, we estimated the highest probability of transmission amongst older individuals aged 50 – 70 years old (Figure 2B). However, this is also likely to reflect symptomaticity by age with symptomatic and severe infections increasing with age and thus more likely to be tested and reported [32, 33]. Accounting for the varying clinical progression by age, with younger age groups less likely to report severe respiratory symptoms or pneumonia (Figure 3), is critical for symptom-based screening and for refining case definitions especially for a novel pathogen. Equally the consistent finding of early onset of fever, cough, and fatigue across all age groups is informative for self-isolation guidance when testing capacity may be limited at the beginning of an epidemic.

There are a number of limitations to our study. Whilst publicly available case reports were initially highly detailed, less information was released as the epidemic progressed. Data on date of symptom onset which we used to estimate *R*_*t*_ was missing for 30% of reported cases. However, the majority of cases with missing onset dates were reported after the end of our trusted period on 23 March so would not have affected our estimates of transmissibility. There was limited information on the age distribution of cases or symptoms especially for large clusters such as the care home outbreak in Chiba prefecture or the live music venues in Osaka. Similarly, the information released varied substantially by prefecture with Tokyo, which was the worst affected, only releasing minimal information beyond age group and sex. This may have biased our estimates of the age-dependent probability of transmission especially for the younger and older age groups associated with these clusters. Finally, we did not follow the entire clinical progression for patients with symptom data.

Therefore, patients may have later developed more severe symptoms that were not presented here. However, this should not affect the variation in symptom type at the time of symptom onset across age groups.

## Conclusions

Detailed outbreak investigation and contact tracing data early in the epidemic was instrumental in guiding Japan’s response to COVID-19. Determining the key characteristics and the natural history of novel pathogens is crucial for setting and implementing successful control measures to avoid within-country or global spread. The data analysed here were the result of robust and timely investigations and demonstrates the improvements to epidemic control as a result of such surveillance. Retrospective analyses of early pandemic responses can provide important insights for future preparedness plans.

## Data Availability

The data used in this analysis were collated from publicly available Japanese Ministry of Health case reports and all urls are made available on GitHub. The repository can be found here: https://github.com/mrc-ide/early-japan.
The full extracted data used and/or analysed during the current study are available from the corresponding author on reasonable request.

## Additional file 1

### List of Abbreviations

PCR: Polymerase Chain reaction
*R*_*t*_: time-varying reproduction number

## Declarations

### Ethics approval and consent to participate

Not applicable. Secondary analysis of published, publicly available data.

### Consent for publication

Not applicable. Secondary analysis of published, publicly available data.

### Availability of data and materials

The data used in this analysis were collated from publicly available Japanese Ministry of Health case reports and all urls are made available on GitHub. The repository can be found here: https://github.com/mrc-ide/early-japan.

The full extracted data used and/or analysed during the current study are available from the corresponding author on reasonable request.

## Authors’ contributions

NI, KAMG, SB, TDM, GC-D, HJTU, and NMF conceived the study; NI collected, translated, and extracted the data; NI, KAMG, SB, TDM, GC-D, and HJTU carried out the analysis; NI, KAMG, SB, and TDM wrote the first draft of the manuscript; all authors contributed to, read, and approved the final draft.

## Acknowledgements

Not applicable

