## Additional material 1 for "COVID-19 in Japan: insights from the first three months of the epidemic"

### *Results*

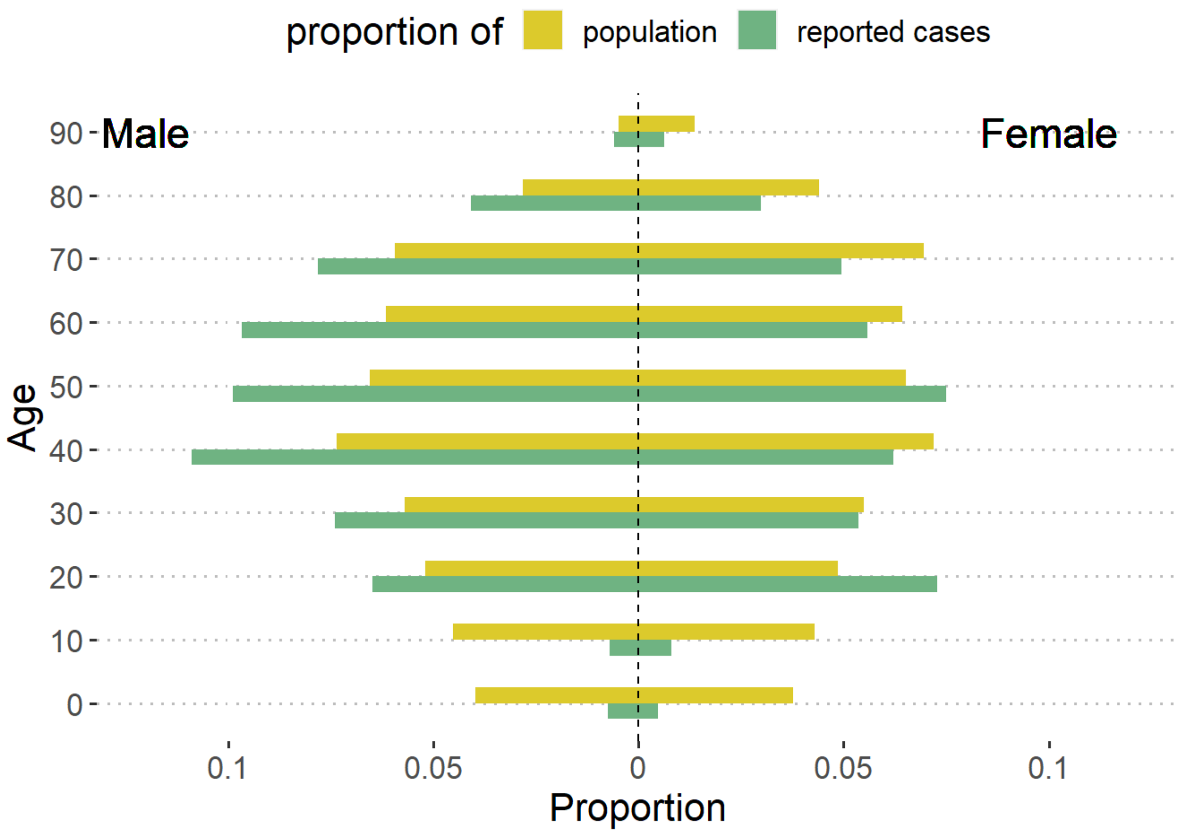


*Supplementary Figure 1: Proportion of the population (yellow) of Japan and confirmed COVID-19 cases (green) by sex and 10-year age bands.*

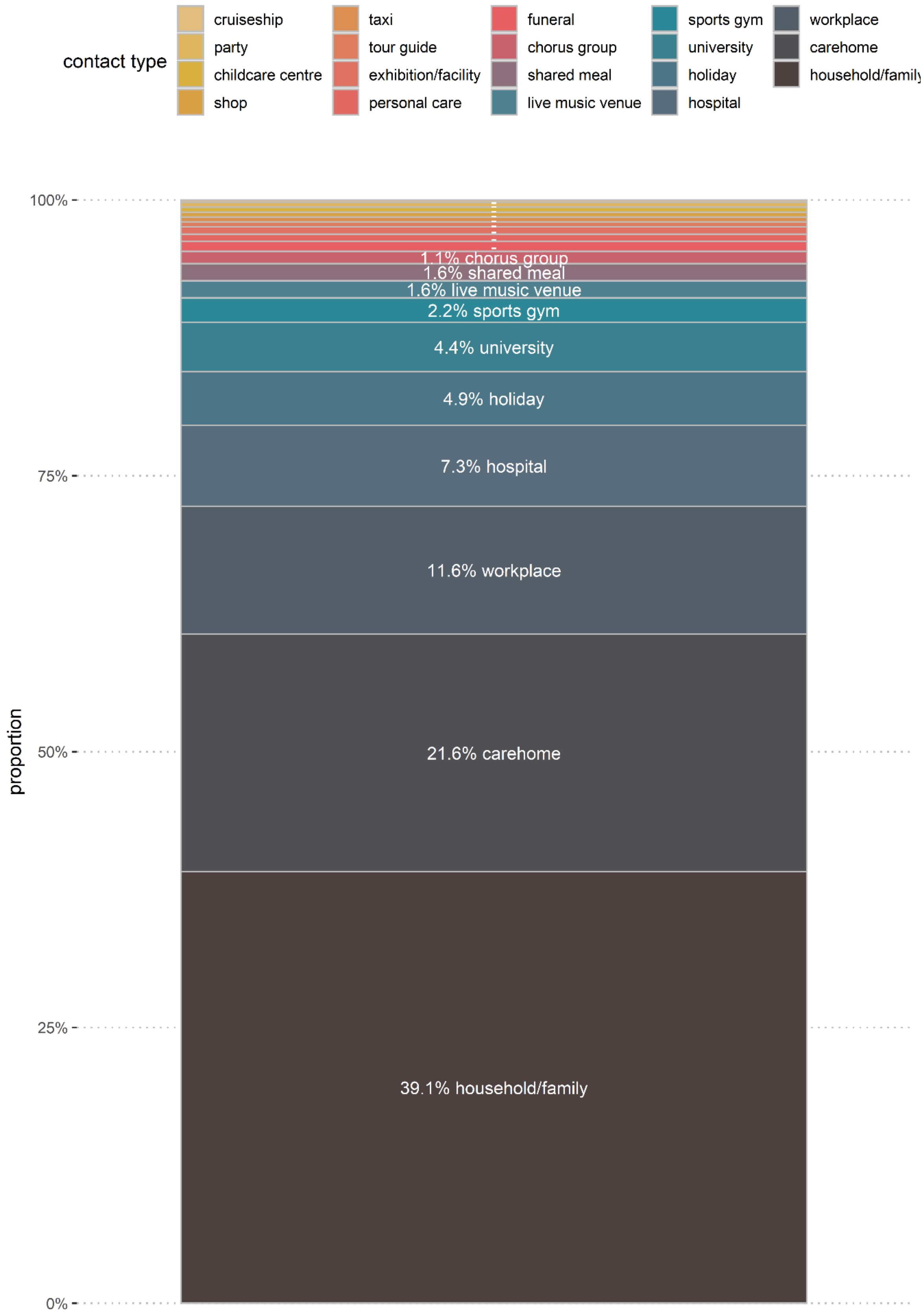


*Supplementary Figure 2: Proportion of known contacts by contact type.*

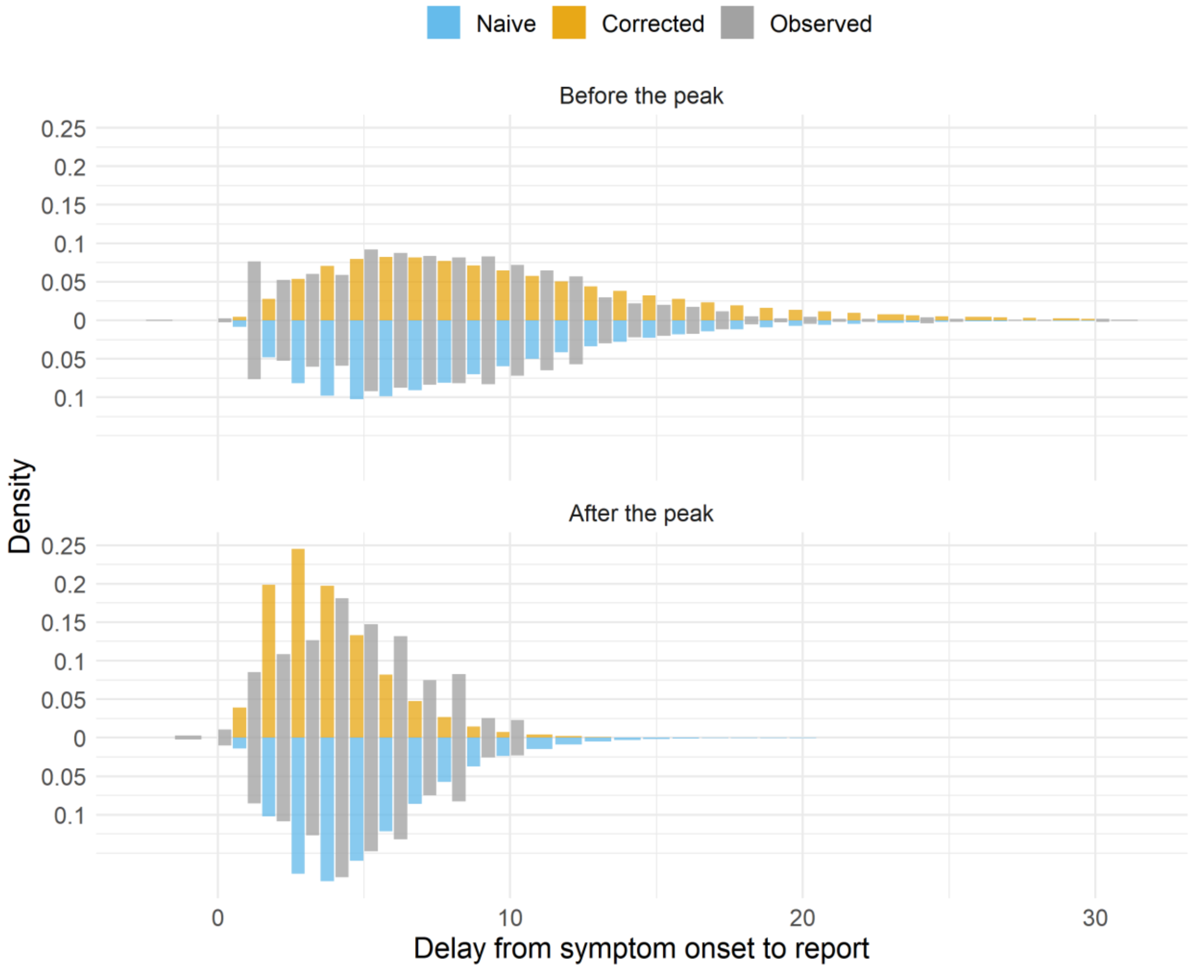


*Supplementary Figure 3: Time between symptom onset and case report for before (top panel) and after (bottom panel) the estimated peak (19 March 2020). Grey bars show the observed delay, the blue and orange bars show the best-fit gamma probability density function for the (blue) naïve delay and (orange) the delay corrected for epidemic growth and decay.*

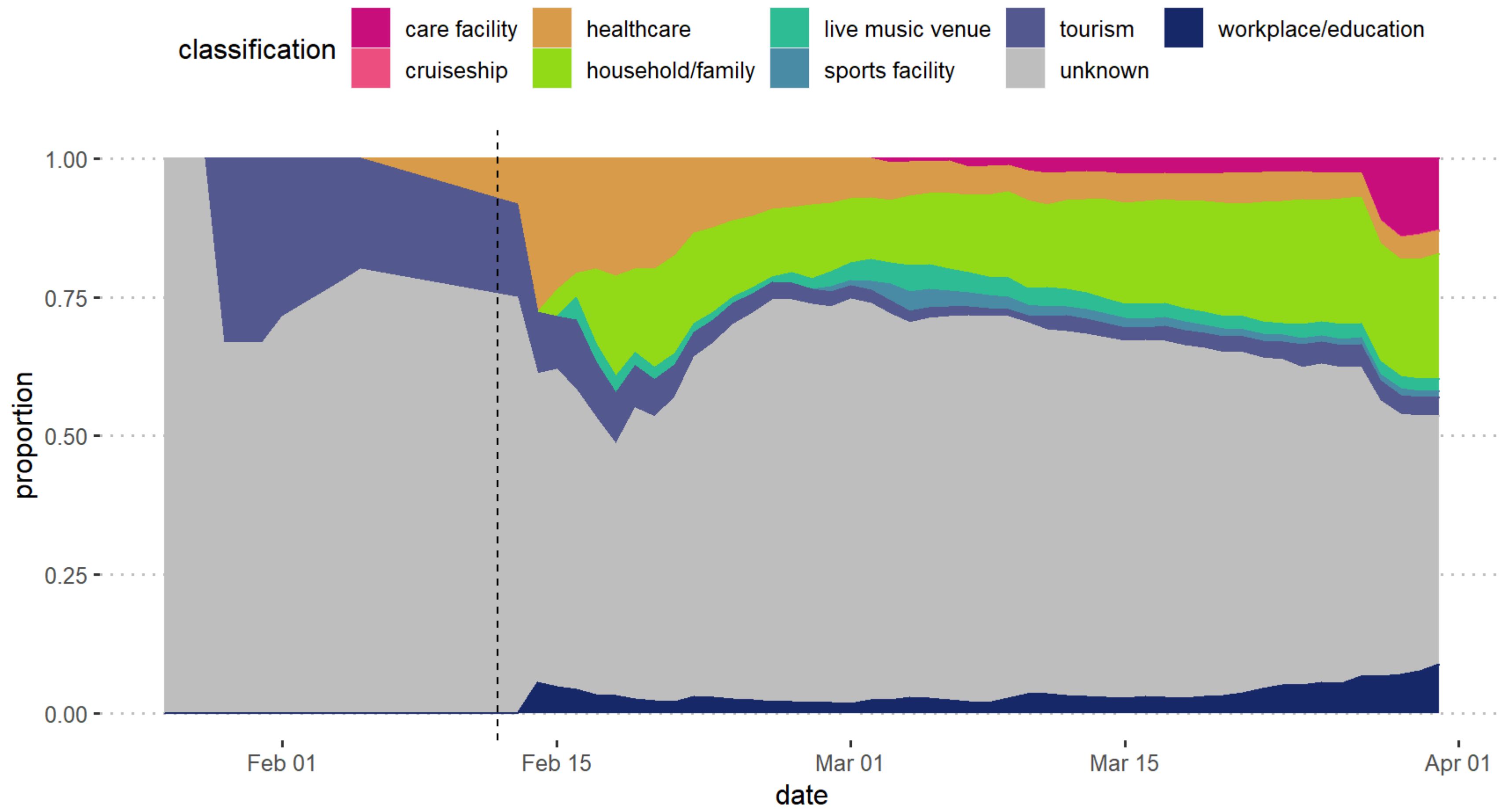


*Supplementary Figure 4: Change in contact exposure types (known and unknown) over time from 14 January to 31 March 2020.*

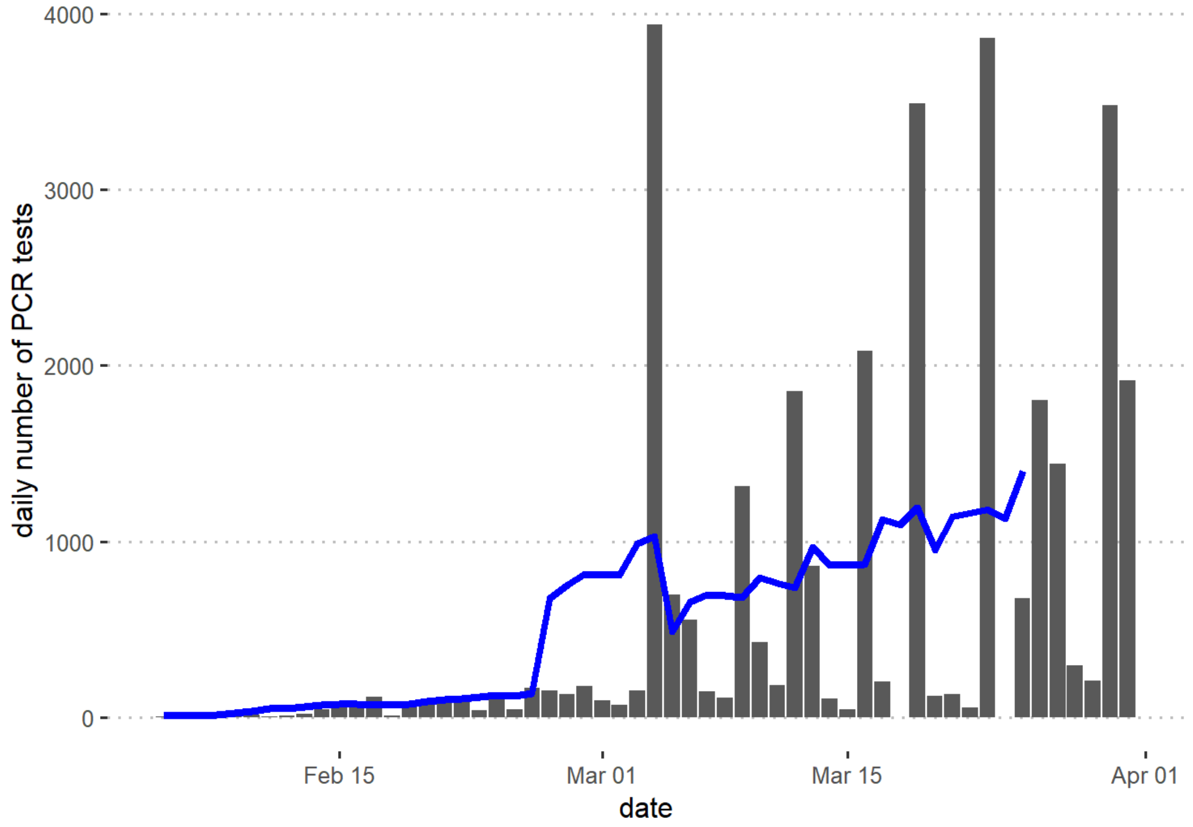


*Supplementary Figure 5: Daily number of PCR tests conducted nationally from 5 Feb to 31 March 2020 as reported by the Ministry of Health Japan* [1].

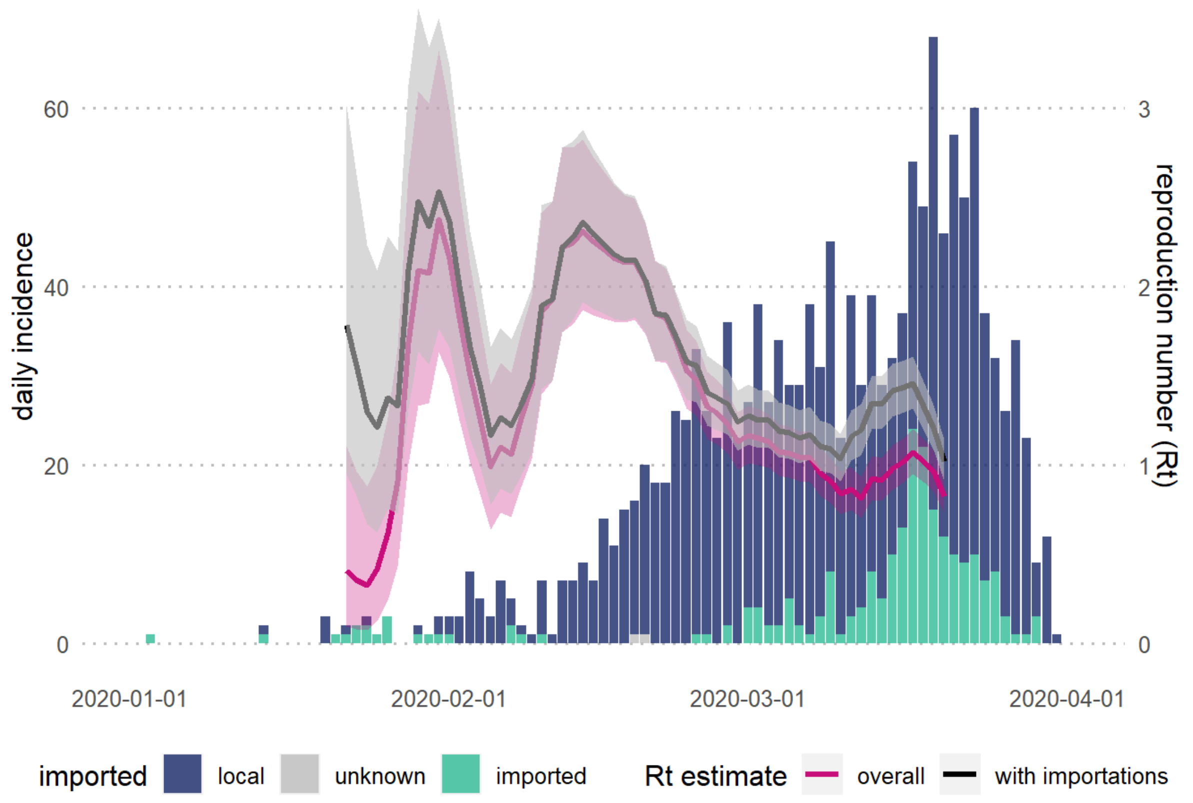


*Supplementary Figure 6: Daily incidence of confirmed COVID-19 cases by date of symptom onset and the estimated time-varying reproduction number ($R_t$) estimated from incidence data truncated using the empirical mean onset-to-reporting delay: overall (pink) and accounting for imported cases (black). The solid line shows the median and the shaded area the 95% credible interval.*

*Supplementary Table 1: Summary of national and regional COVID-19 interventions up to 31 March 2020*

| region | Date | classification | intervention | url |
|---|---|---|---|---|
| **national** | 2020/01/27 | change in legal scope | PM designates COVID as infectious disease under infectious diseases control law | http://www.asahi.com/ajw/articles/AJ202001270041.html |
| **national** | 2020/01/27 | change in legal scope | PM designates COVID as quarantinable infectious disease under the Quarantine Act | http://www.asahi.com/ajw/articles/AJ202001270041.html |
| **national** | 2020/01/29 | repatriation | start of repatriation of Japanese citizens from Hubei province | http://japan.kantei.go.jp/98_abe/actions/202001/_00037.html |
| **national** | 2020/01/30 | expert meeting | Japan Anti-Coronavirus National Task Force established | https://japan.kantei.go.jp/98_abe/actions/202001/_00034.html |
| **national** | 2020/02/01 | travel restrictions | restrictions on foreign citizens who had visited Hubei province within 14 days | https://japan.kantei.go.jp/98_abe/actions/202002/_00001.html |
| **national** | 2020/02/01 | travel restrictions | restrictions on those with a Chinese passport issued from Hubei province | https://japan.kantei.go.jp/98_abe/actions/202002/_00001.html |
| **national** | 2020/02/01 | travel restrictions | restrictions on foreign citizens who had visited Zheijang province within 14 days | https://asia.nikkei.com/Spotlight/Coronavirus/Japan-expands-entry-restrictions-to-virus-hit-Zhejiang |
| **national** | 2020/02/01 | travel restrictions | restrictions on those with a Chinese passport issued from Zheijang province | https://asia.nikkei.com/Spotlight/Coronavirus/Japan-expands-entry-restrictions-to-virus-hit-Zhejiang |
| **national** | 2020/02/01 | emergency investment in healthcare | MOH instructed municipal and prefectural governments to establish specialised COVID-19 centres and outpatient wards | https://mainichi.jp/articles/20200203/k00/00m/040/227000c |
| **Kanagawa** | 2020/02/05 | quarantine | placed the Diamond Princess under quarantine in Yokohama | http://japan.kantei.go.jp/98_abe/actions/202002/_00006.html |
| **national** | 2020/02/05 | testing policy | begun preparations to strengthen testing capacity in all prefectures | http://japan.kantei.go.jp/98_abe/actions/202002/_00006.html |
| **national** | 2020/02/06 | travel restrictions | denied entry to the MS Westerdam from Hong Kong | http://japan.kantei.go.jp/98_abe/actions/202002/_00010.html |
| **national** | 2020/02/12 | testing policy | expanded testing criteria to all those with symptoms regardless of travel history | http://japan.kantei.go.jp/98_abe/actions/202002/_00016.html |
| **national** | 2020/02/12 | financial support | financial support for small-medium businesses affected by the outbreak | http://japan.kantei.go.jp/98_abe/actions/202002/_00019.html |
| **national** | 2020/02/14 | emergency investment in healthcare | introduced coronovirus consultation system -- this was specialised covid medical services | http://japan.kantei.go.jp/98_abe/actions/202002/_00024.html |
| **national** | 2020/02/16 | expert meeting | first Novel Coronavirus Expert Meeting | https://www.kantei.go.jp/jp/singi/novel_coronavirus/senmonkakaigi/konkyo.pdf |
| **national** | 2020/02/17 | repatriation | end of repatriation of Japanese citizens from Hubei province | http://japan.kantei.go.jp/98_abe/actions/202001/_00037.html |
| **national** | 2020/02/17 | public information campaign | released national guidelines on COVID-19 testing | https://www.mhlw.go.jp/content/10900000/000607629.pdf |
| **national** | 2020/02/24 | expert meeting | president convened ministerial meeting in response to confirmed human-to-human transmission | http://japan.kantei.go.jp/98_abe/actions/202001/_00024.html |

| | | | | |
|---|---|---|---|---|
| **national** | 2020/02/24 | expert meeting | second Novel Coronavirus Expert Meeting | https://www.nippon.com/en/news/yjj2020022400554/japanese-experts-discuss-basic-coronavirus-policy.html |
| **national** | 2020/02/25 | combined | | |
| **national** | 2020/02/25 | public information campaign | introduction of the basic policies for novel coronavirus disease control | https://www.kantei.go.jp/jp/singi/novel_coronavirus/th_siryou/kihonhousin.pdf |
| **national** | 2020/02/25 | cancel public events | request for the suspension of large-scale gatherings | https://www.mhlw.go.jp/content/10200000/000603611.pdf |
| **national** | 2020/02/25 | stay home request | asked all those with cold symptoms to stay home | https://www.nippon.com/en/news/l00269/coronavirus-basic-policy-impacts-japan%e2%80%99s-health-education-systems.html?cx_recs_click=true |
| **national** | 2020/02/25 | contact tracing | established the Cluster Response Team to identify small clusters of infections | https://www.mhlw.go.jp/stf/newpage_09743.html |
| **Hokkaido** | 2020/02/26 | school closure | closure of elementary, junior high and high schools | https://english.kyodonews.net/news/2020/02/b95d548f279f-hokkaido-eyes-temporary-shutdown-of-public-schools-to-fight-virus.html |
| **national** | 2020/02/27 | emergency investment in healthcare | expand national health insurance to cover COVID-19 testing | https://www.nippon.com/en/news/ntv20200227002/coronavirus-national-health-insurance-to-cover-virus-test.html?cx_recs_click=true |
| **Hokkaido** | 2020/02/28 | state of emergency | declaration of a new coronavirus emergency | https://www.pref.hokkaido.lg.jp/ss/tkk/koronaseng en.html |
| **national** | 2020/03/02 | school closure | closure of elementary, junior high and high schools | https://english.kyodonews.net/news/2020/02/c3c57bbce11d-breaking-news-govt-will-ask-all-schools-in-japan-to-shut-for-virus-fears-abe.html |
| **national** | 2020/03/09 | expert meeting | reconvened the expert meeting | https://www.mhlw.go.jp/content/10900000/000608425.pdf |
| **national** | 2020/03/26 | expert meeting | reconvened the expert meeting | https://www.mhlw.go.jp/content/10900000/000620826.pdf |
| **Tokyo** | 2020/03/30 | stay home request | request residents to refrain from non-essential outings for two weeks | https://japantoday.com/category/national/Koike-calls-for-fewer-outings-says-state-of-emergency-up-to-PM |

*Supplementary Table 2: Age-specific transmission probability (95% CI, 3 d.p) matrix for all provinces in the first wave of the epidemic in Japan (N=1276). The probability is defined as the frequency of reported transmission events from the infector age-group to the infectee age-group divided by the total number of linked transmission pairs*

| | **Infector** | | | | | | | | | |
|---|---|---|---|---|---|---|---|---|---|---|
| **Infectee** | **0-9** | **10-19** | **20-29** | **30-39** | **40-49** | **50-59** | **60-69** | **70-79** | **80-89** | **90+** |
| **0-9** | 0 (0-0) | 0.002 (0.002-0.006) | 0 (0-0) | 0.006 (0.002-0.013) | 0.003 (0-0.008) | 0.005 (0-0.011) | 0 (0-0) | 0 (0-0) | 0 (0-0) | 0 (0-0) |
| **10-19** | 0 (0-0) | 0 (0-0) | 0.002 (0-0.005) | 0.002 (0-0.005) | 0.003 (0-0.008) | 0.003 (0-0.008) | 0 (0-0) | 0.002 (0-0.005) | 0 (0-0) | 0 (0-0) |
| **20-29** | 0 (0-0) | 0 (0-0) | 0.034 (0.021-0.049) | 0.006 (0-0.013) | 0.005 (0-0.011) | 0.026 (0.013-0.039) | 0.015 (0.006-0.024) | 0.005 (0-0.011) | 0.006 (0.002-0.013) | 0 (0-0) |
| **30-39** | 0.002 (0.002-0.006) | 0 (0-0) | 0.005 (0-0.011) | 0.015 (0.006-0.024) | 0.006 (0-0.013) | 0.006 (0.002-0.013) | 0.013 (0.005-0.023) | 0 (0-0) | 0.002 (0-0.005) | 0 (0-0) |
| **40-49** | 0 (0-0) | 0 (0-0) | 0.01 (0.003-0.018) | 0.018 (0.008-0.029) | 0.034 (0.021-0.05) | 0.026 (0.015-0.041) | 0.011 (0.003-0.021) | 0.015 (0.006-0.026) | 0.01 (0.003-0.018) | 0 (0-0) |
| **50-59** | 0 (0-0) | 0 (0-0) | 0.008 (0.002-0.015) | 0.01 (0.003-0.018) | 0.024 (0.013-0.036) | 0.073 (0.055-0.094) | 0.041 (0.026-0.058) | 0.024 (0.015-0.037) | 0.023 (0.011-0.036) | 0.006 (0.002-0.013) |
| **60-69** | 0 (0-0) | 0 (0-0) | 0.01 (0.003-0.018) | 0.01 (0.003-0.019) | 0.013 (0.005-0.023) | 0.047 (0.031-0.065) | 0.065 (0.047-0.086) | 0.042 (0.028-0.058) | 0.023 (0.011-0.037) | 0.011 (0.003-0.019) |
| **70-79** | 0 (0-0) | 0 (0-0) | 0 (0-0) | 0.003 (0-0.008) | 0.013 (0.005-0.023) | 0.019 (0.01-0.031) | 0.029 (0.016-0.042) | 0.060 (0.042-0.078) | 0.021 (0.011-0.032) | 0.002 (0-0.005) |
| **80-89** | 0 (0-0) | 0 (0-0) | 0 (0-0) | 0 (0-0) | 0.002 (0-0.005) | 0.019 (0.01-0.032) | 0.016 (0.006-0.026) | 0.013 (0.005-0.023) | 0.034 (0.023-0.05) | 0.003 (0-0.008) |
| **90+** | 0 (0-0) | 0 (0-0) | 0 (0-0) | 0 (0-0) | 0.003 (0-0.008) | 0.013 (0.005-0.023) | 0.011 (0.003-0.021) | 0.005 (0-0.011) | 0.002 (0-0.005) | 0.003 (0-0.008) |

The onset of pneumonia varies most significantly between youngest and oldest age groups (Supplementary Table 3). We also estimate significant differences in the onset of cough between 20-29 years and 60-69 or 80+ years and between 50-59 years and 80+ years; the onset of fatigue between 0-19 years and 20-29 or 60-69 years and between 50-59yo and 80+yo; and the onset of respiratory symptoms between 50-59 and 80+ years.

*Supplementary Table 3: Summary of p-values of the Kolmogorov-Smirnov statistic to assess whether samples of time from symptom onset to first occurrence of a specific symptom from different age groups are drawn from the same distribution. Only rows where a significant p-value appears are shown with minimum p-value per row shown in the "minimum p-value" column. Significance symbols are as follows: * <0.05, ** <0.01, *** <0.001 and significant results are highlighted in blue.*

| **Symptom** | | | | | **Ages** | | | |
|---|---|---|---|---|---|---|---|---|
| **Cough** | **Fatigue** | **Fever** | **Pneumonia** | **Respiratory** | **Age group 1** | **Age group 2** | **Minimum p-value** | **Significance** |
| **0.46** | 0.02 | 0.6 | 0.56 | 0.99 | 0-19 | 20-29 | 0.02 | * |
| **0.8** | 0.08 | 0.58 | 0.02 | 0.99 | 0-19 | 50-59 | 0.02 | * |
| **0.96** | 0.04 | 0.84 | 0.28 | 0.61 | 0-19 | 60-69 | 0.04 | * |
| **0.87** | 0.11 | 0.85 | 0.02 | 0.86 | 0-19 | 70-79 | 0.02 | * |
| **0.25** | 0.47 | 0.6 | 0 | 0.61 | 20-29 | 40-49 | 0 | ** |
| **0.76** | 0.91 | 1 | 0 | 0.9 | 20-29 | 50-59 | 0 | ** |
| 0.02 | 0.89 | 0.98 | 0.15 | 0.44 | 20-29 | 60-69 | 0.02 | * |
| **0.38** | 0.75 | 1 | 0 | 0.83 | 20-29 | 70-79 | 0 | *** |
| 0 | 0.14 | 0.65 | 0.04 | 0.1 | 20-29 | 80+ | 0 | ** |
| **0.99** | 0.12 | 0.99 | 0.05 | 0.27 | 30-39 | 50-59 | 0.05 | * |
| **0.21** | 0.74 | 0.87 | 0.03 | 0.06 | 50-59 | 60-69 | 0.03 | * |
| 0.03 | 0.03 | 0.28 | 0.29 | 0.03 | 50-59 | 80+ | 0.03 | * |
| **0.47** | **0.94** | **1** | 0.04 | **0.75** | **60-69** | **70-79** | **0.04** | * |
